# Weaker functional segregation of the sensory and default mode networks in trauma-related intrusive memories

**DOI:** 10.64898/2026.09.18.26363166

**Authors:** Kevin J. Clancy, Boyu Ren, Isabelle M. Rosso

**Affiliations:** Division of Depression and Anxiety Disorders, McLean Hospital, Belmont, MA; Department of Psychiatry, Harvard Medical School, Boston, MA; Laboratory for Psychiatric Biostatistics, McLean Hospital, Belmont, MA

**Author notes:** **Corresponding Author:** Kevin J. Clancy, Ph.D., **Mailing Address:** 115 Mill St., Belmont, MA 02478.

## Abstract

Trauma-related intrusive memories (TRIMs) are a core symptom of posttraumatic stress disorder and cause significant distress and impairment for trauma-exposed individuals due to their vivid sensory features that intrude on ongoing thought. These phenomenological properties point to a neuropathology that spans autobiographical memory, spontaneous thought, and sensory perception. The hierarchical organization of the cortex maximally segregates the default mode network (DMN), which supports autobiographical memory and ongoing thought, from the sensory cortices (SC). Accruing evidence implicates altered interactions between the DMN and SC in PTSD and TRIMs, suggesting a breakdown in this segregation. We sought to test whether diminished DMN-SC segregation, indexed by a shorter functional distance between the DMN and SC within cortical connectivity gradients, was associated with the vividness and intrusiveness of TRIMs. One-hundred eight trauma-exposed adults completed two weeks of ecological momentary assessments (EMAs) of TRIM properties followed by resting-state fMRI. Gradient analysis of cortical functional connectivity was performed to compute the Euclidean distance between resting-state networks. Pearson correlations revealed an association between a shorter DMN-SC distance and greater TRIM vividness and intrusiveness, with multivariate models indicating a specificity for intrusiveness. Importantly, the DMN-SC distance was not associated with clinician-rated symptom severity and explained unique variance above and beyond conventional functional connectivity measures. These findings suggest that a loss of functional segregation between the DMN and SC – a core organizational feature of the cortical hierarchy – is associated with the intrusive sensory vividness of TRIMs, offering insights into candidate mechanisms of and neurotherapeutic targets for intrusive trauma memories.

## INTRODUCTION

Intrusive memories of a traumatic experience are prevalent among individuals exposed to trauma and are leading predictors of the onset, persistence, and severity of post-traumatic psychiatric sequelae, including posttraumatic stress disorder (PTSD), major depressive disorder, and substance use disorder [1–3]. Independently, these memories cause clinically significant distress and impairment, regardless of psychiatric diagnosis [4]. As such, trauma-related intrusive memories (TRIMs) represent critical intervention targets for trauma-exposed individuals, underscoring the need to better characterize their underlying neural correlates to develop mechanism-based therapeutics.

Intrusive memories are a common psychological phenomenon that arise as part of normal autobiographical memory processes. However, unlike general intrusive memories, TRIMs often cause significant distress and impairment due to their vivid sensory detail that intrudes on ongoing cognition, with marked difficulties to suppress or control the intruding memory [5–9]. Their vividness and intrusiveness can contribute to functional impairment through “attentional hijacking” and urgent, often iatrogenic, attempts to dampen their vividness [10]. Evidence furthers suggests that these intruding sensory-rich properties may distinguish TRIMs from other forms of autobiographical memory [11], highlighting their clinical significance. Moreover, these phenomenological reports of TRIM properties provide insights into potential neural mechanisms, hinting towards a pathophysiology that spans the neural systems supporting sensory perception and autobiographical memory.

Evidence from neuroimaging studies indicates dysfunction of the neural systems supporting sensory perception and autobiographical memory in trauma-exposed individuals and patients with PTSD. With regards to sensory perception, longstanding evidence demonstrates robust deficits in sensory gating across sensory modalities in PTSD [12–16], with more recent evidence pointing towards exaggerated activation, or disinhibition, within the sensory cortices (SC) of PTSD patients [17, 18]. Alterations in the structure and function of the visual and somatosensory cortices have been increasingly implicated in the development and maintenance of posttraumatic stress symptoms, particularly reexperiencing symptoms [19–23]. With regards to autobiographical memory, trauma exposure and PTSD are associated with reduced activity and connectivity within the default mode network (DMN), which supports autobiographical memory [24–29]. Moreover, there is emerging evidence that links these dysfunctions in trauma-exposed individuals, revealing exaggerated coactivation and altered connectivity between the SC and DMN as well as deficits in inhibitory neural activity across these systems [23, 30, 31]. Taken together, these findings converge on a pattern of hyperactive SC and hypoactive DMN and aberrant interactions between them.

This emergent pattern contrasts with the intrinsic dynamics and organization of the human brain. At rest, activity within the DMN is dominant while the SC is deactivated, reflecting an intrinsic anticorrelation between these networks [32, 33]. The functional organization of the cortex is structured in a way that mirrors this anticorrelation, maximally segregating the SC from the DMN [34]. This organization is reflected in patterns of cortical functional connectivity, whereby dimensionality reduction techniques of cortex-wide functional connectivity reveal organizational gradients based on similarity of connectivity patterns [35]. The principal connectivity gradient, which accounts for the greatest variance in cortical functional connectivity, is anchored at one end by unimodal sensory (visual and sensorimotor) cortices, which support basic sensory perception. The other end of the principal gradient is anchored by transmodal regions of the DMN, which support the highest levels of cognition like autobiographical memory and a sense of self in time and place (i.e., autonoetic consciousness) [36]. This maximal segregation of the SC and DMN along the principal gradient reflects their highly differentiated functional connectivity profiles. This intrinsic segregation of external- sensory and internal-self-referential systems constitutes a core feature of the cortical hierarchy, which is believed to support adaptive cognition and protect higher-order cognitive networks from spontaneous sensory intrusions [34, 35, 37].

Linking these lines of evidence, we sought to examine the functional segregation of the SC and DMN along the gradient of functional connectivity in trauma-exposed adults and its association with the phenomenological properties of TRIMs. Combining daily ecological momentary assessments (EMAs) of TRIM properties with functional imaging of resting-state connectivity in trauma-exposed adults, we tested the hypothesis that a shorter functional distance between the SC and DMN within the gradients of functional connectivity, reflecting lower network segregation and a compression of the cortical hierarchy, would be associated with the vividness and intrusiveness of TRIMs.

## METHODS

### Participants and Procedures

We enrolled 133 adults (18-65 years old) from the community who reported exposure to a Criterion A traumatic event and endorsed at least two TRIMs per week, as part of a larger study [31, 38–40]. Study procedures were approved by the Mass General Brigham Human Research Committee, and all participants provided written informed consent. Inclusion criteria consisted of: exposure to a DSM-5 Criterion A trauma and currently (past month) experiencing at least two TRIMs per week. Exclusion criteria included: left-handedness, medical conditions that could confound results, a history of moderate to severe traumatic brain injury, current (past month) moderate-to-severe alcohol or substance use disorder, and current psychotic disorder or manic episode.

Participants completed daily ecological momentary assessments (EMAs) of TRIMs for two weeks, followed by a neuroimaging visit that included the Clinician-Administered PTSD Scale for DSM-5 (CAPS-5) [41] and resting-state functional magnetic resonance imaging (rs-fMRI). rs-fMRI data was excluded due to confounds or poor quality: falling asleep (n = 2), artifacts or anatomical defects (n = 3), and excessive motion (detailed below; n = 12). Per eligibility requirements, participants who reported fewer than two TR-IMs per week were excluded from analyses (n = 8), resulting in a final analyzed sample of 108 participants.

### Clinician-Administered PTSD Scale for DSM-5 (CAPS-5)

The CAPS-5 [41] was administered by doctoral-level clinicians to determine a PTSD diagnosis and assess PTSD symptom severity. The CAPS-5 consists of 20 symptoms of PTSD and are rated on frequency and intensity to determine a symptom severity score (Likert scale: 0 = absent to 4 = Extreme). A CAPS-5 total score was calculated as the sum of all 20 symptoms, and the intrusive reexperiencing symptom cluster (Cluster B) score as the sum of scores for the five symptoms in that cluster (B1-B5). Table 1 shows that most participants met diagnostic criteria for PTSD in this sample.

**Table 1.** Demographic and clinical characteristics. Mean ± standard deviation or N (%).

| Total Sample ($n = 108$) | |
| --- | --- |
| Age (years) | 32.2 ± 10.2 |
| Sex assigned at birth |  |
| Female | 86 (80%) |
| Male | 22 (20%) |
| Gender |  |
| Woman | 74 (69%) |
| Man | 22 (20%) |
| Gender fluid/non-binary | 12 (11%) |
| Race |  |
| White | 72 (66%) |
| Multiracial | 21 (19%) |
| Asian | 6 (6%) |
| Black | 5 (5%) |
| Unknown or Not Reported | 4 (4%) |
| Ethnicity |  |
| Non-Hispanic | 101 (94%) |
| Hispanic | 7 (6%) |
| Current Psychotropic Medication | 53 (49%) |
| Antipsychotic* | 6 (6%) |
| Antidepressant | 38 (35%) |
| Stimulant | 5 (5%) |
| Mood Stabilizer | 3 (3%) |
| Sedative/Hypnotic | 2 (2%) |
| CAPS-5 Cluster B Severity (Reexperiencing Symptoms) | 9.9 ± 3.4 |
| CAPS-5 Total Score | 34.9 ± 10.9 |
| CAPS-5 PTSD Diagnosis | 87 (81%) |

**Table 1.** Demographic and clinical characteristics. Mean ± standard deviation or N (%).
| Total Sample (n = 108) |  |
| --- | --- |
| Age (years) | 32.2 ± 10.2 |
| Sex assigned at birth |  |
| Female | 86 (80%) |
| Male | 22 (20%) |
| Gender |  |
| Woman | 74 (69%) |
| Man | 22 (20%) |
| Gender fluid/non-binary | 12 (11%) |
| Race |  |
| White | 72 (66%) |
| Multiracial | 21 (19%) |
| Asian | 6 (6%) |
| Black | 5 (5%) |
| Unknown or Not Reported | 4 (4%) |
| Ethnicity |  |
| Non-Hispanic | 101 (94%) |
| Hispanic | 7 (6%) |
| Current Psychotropic Medication | 53 (49%) |
| Antipsychotic* | 6 (6%) |
| Antidepressant | 38 (35%) |
| Stimulant | 5 (5%) |
| Mood Stabilizer | 3 (3%) |
| Sedative/Hypnotic | 2 (2%) |
| CAPS-5 Cluster B Severity (Reexperiencing Symptoms) | 9.9 ± 3.4 |
| CAPS-5 Total Score | 34.9 ± 10.9 |
| CAPS-5 PTSD Diagnosis | 87 (81%) |

### Ecological momentary assessments

Participants completed two-weeks of EMAs assessing the phenomenological properties of TRIMs. This consisted of 3 daily surveys delivered on a semi-random schedule via the MetricWire smartphone app. Surveys consisted of a query about the number of TRIMs experienced since the last survey, followed by 18 prompts assessing TRIM properties. These prompts were adapted from the Autobiographical Memory Questionnaire (AMQ) and rated on a 0-4 Likert scale [9, 42]. Ratings for each property were averaged across all completed surveys and grouped into vividness, visual detail, reliving, emotional intensity, fragmentation, and intrusiveness for each participant [43].

### Functional magnetic resonance imaging

Eyes-open resting-state fMRI data (13 minutes, 976 volumes) were acquired on a 3T Siemens Prisma scanner with a 64-channel head coil using the Human Connectome Project (HCP) Lifespan protocol (T2-weighted echoplanar images; TR/TE: 800/37 ms, in-plane resolution: 2mm; voxels: 2mm isotropic; multiband factor = 8; anterior-posterior phase encoding; one run of 976 frames, ∼13 minutes in length) [44]. In addition, T1-weighted 3D MPRAGE anatomical images were obtained using the HCP 0.8mm resolution sequence (TR/TEs: 2500/1.81/3.6/5.39/7.18; flip angle: 8 deg; FOV: 256 x 240; voxel size: 0.8mm isotropic). MRI data were preprocessed using fMRIPrep version 20.2.7 [45], followed by additional denoising of white matter and cerebrospinal fluid signals [46], scrubbing of motion outliers (framewise displacement > 0.5 mm) [47]; and high pass (0.01 Hz) filtering using CONN toolbox scripts [48]. Participants were excluded if their mean framewise displacement (FD) exceeded 0.5 mm or greater than 20% of volumes exceeded FD = 0.5mm (n = 10) [49].

rs-fMRI were segmented into 400 cortical parcels using the Schaefer atlas [50]. Cleaned timeseries for each parcel were submitted to pairwise Pearson’s correlation analyses, resulting in a 400×400 connectivity matrix. The resulting connectivity matrix was submitted to functional connectivity gradient analyses.

### Functional connectivity gradient analysis

Connectivity gradient analyses were performed using the BrainSpace toolbox in Matlab [51]. A normalized angle similarity matrix was generated from a sparsified (10%) 400×400 cortex-wide connectivity matrix and submitted to diffusion map embedding to compute cortical connectivity gradients. These gradients reflect one-dimensional patterns explaining variance in functional connectivity across the 400 cortical parcels. The absolute gradient value indicates how strongly each parcel’s connectivity resembles one of the two primary connectivity patterns at the gradient extremes, while the sign indicates which pattern it most closely resembles. Parcels with similar connectivity patterns have similar gradient values.

To allow for between-subjects comparisons, gradients for each participant were aligned to HCP template gradients from the BrainSpace toolbox using Procrustes alignment [51, 52]. A scree-plot determined that two gradients explained the most meaningful amount of variance in cortical connectivity (Figure 1A). Consistent with prior work [35], the first, or principal, gradient was characterized by the SC (visual and somatomotor networks) at one end and the DMN at the other end. The secondary gradient was characterized by the visual and somatomotor networks at opposing ends with the DMN situated in between them (Figure 1B). These two gradients can be plotted in a two-dimensional Euclidean space to visualize the integration and segregation of individual cortical parcels based on the similarity of their connectivity profiles (Figure 1C).

**Figure 1.**
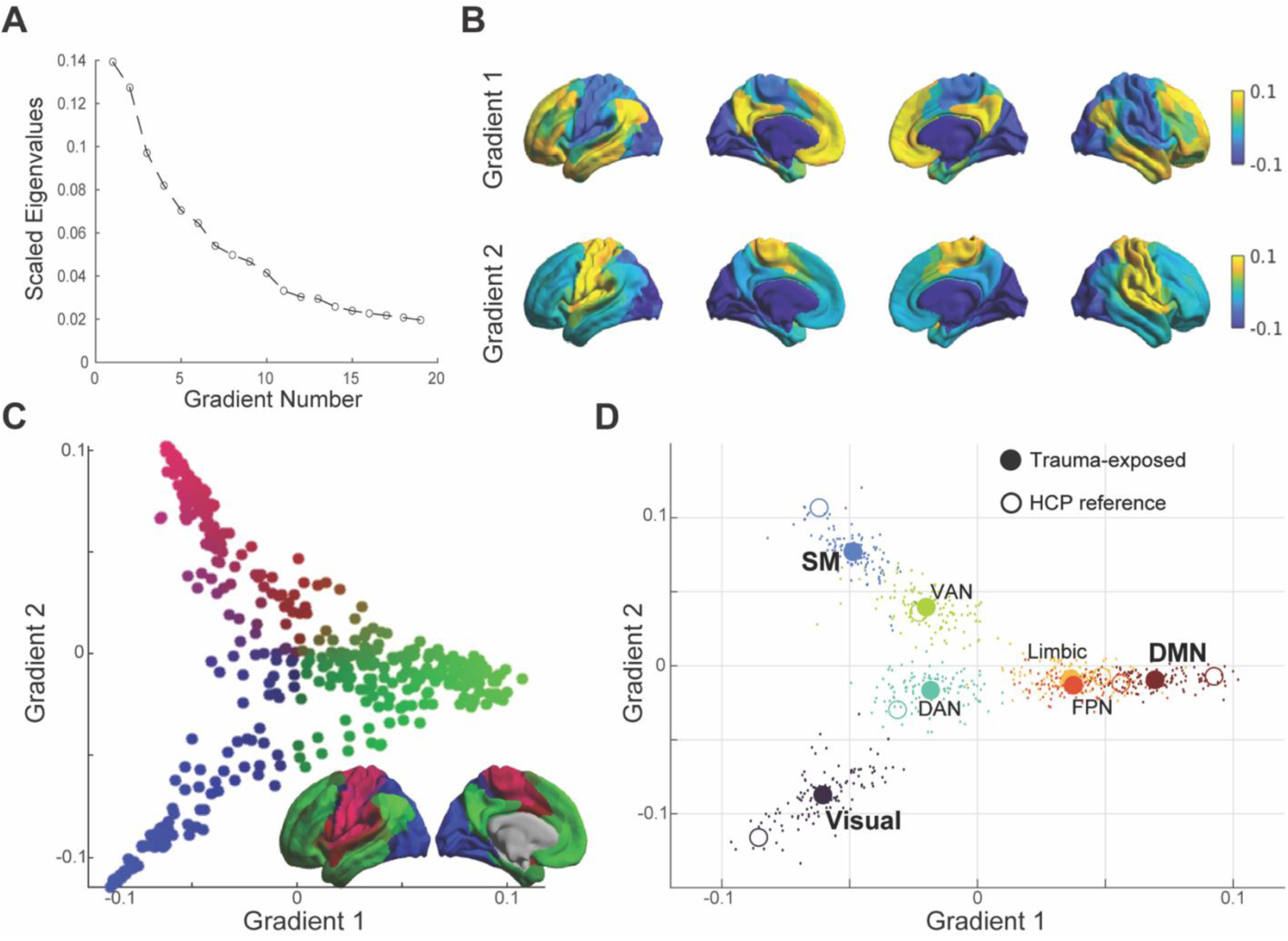
Functional Connectivity Gradients. A) Scree plot depicting the variance explained for each estimated gradient, with a sharp drop in variance explained after Gradient 2. B) The first two gradients projected to the cortical surface (Schaefer 400 parcel atlas). Gradient 1 is characterized by a unimodal sensory cortex to DMN gradient, while Gradient 2 is characterized by a visual to somatosensory network gradient. C) The first two gradients plotted in 2D Euclidean space. Each dot represents an individual cortical parcel, which is colored based on its position within this 2D space and projected onto the cortical surface. Parcels with more similar connectivity patterns have a more similar color. D) Yeo’s 7 major resting-state networks were positioned within this 2D space, color-coded by network. Solid circles represent the group average gradient values for each network within our trauma-exposed sample while individual points represent individual participants’ network gradient values. The open circles represent the reference HCP dataset, demonstrating a notable compression of the Visual, Somatomotor, and Default Mode Networks within the 2D gradient space in our trauma exposed adults. SM = somatomotor network; DAN = dorsal attention network; VAN = ventral attention network; FPN = frontoparietal network; DMN = default mode network.

These cortical gradients were used to index the functional segregation of cortical networks. Cortical parcels were assigned to one of the seven Yeo networks based on the Schaefer-Yeo atlas [50, 53]. At the participant level, principal and secondary gradient values for parcels within each network were averaged together to situate each network within Euclidean space (Figure 1D). Euclidean distance was then calculated between each network to reflect how segregated the networks were, with a larger distance reflecting greater segregation. The present study focused on the distance between the DMN and SC, computed as the average of DMN-Visual and DMN- Somatomotor distances.

### Statistical Analyses

Pearson correlations assessed associations between TRIM properties and DMN-SC Euclidean distance to test whether greater vividness and intrusiveness of TRIMs was associated with less segregation, or a shorter distance, between the DMN and SC. Correction for multiple comparisons across the different TRIM properties was performed using Bonferroni correction (p < 0.05 / 6 TRIM properties = p < 0.008). A multivariate linear regression model incorporating all TRIM properties as predictors of the DMN-SC distance was performed to test for specificity of associations when controlling for their shared variance across all TRIM properties. Sensitivity analyses were performed to examine associations between DMN-SC Euclidean distance and CAPS-5 symptom severity and frequency of TRIMs. Similarly, Welch’s two sample t-test examined the effect of PTSD diagnosis on DMN-SC distance and additional linear regressions examined PTSD diagnosis as a moderator of the associations between DMN-SC Euclidean distance and TRIM properties. Finally, for TRIM properties demonstrating a significant association with DMN-SC distance, control analyses examined whether the DMN-SC Euclidean distance explained unique variability in TRIM properties beyond that accounted for by conventional functional connectivity strength between the DMN and SC. As these control analyses were considered supplemental to our primary, hypothesis-testing analyses, correction for multiple comparisons across the evaluated TRIM properties was not performed.

## RESULTS

### Less DMN-SC segregation was associated with the vividness and intrusiveness of TR-IMs

Correlating the Euclidean distance between the DMN and SC with TRIM properties revealed a significant association between a shorter DMN-SC distance and greater TRIM vividness (*r* = −0.28, *p* = 0.004) and intrusiveness (*r* = −0.26, *p* = 0.006; Figure 2). Additional associations that did not survive correction for multiple comparisons were seen with the visual features (*r* = −0.25, p = 0.010), reliving (*r* = −0.21, *p* = 0.029), and emotional intensity (*r* = −0.19, *p* = 0.049) of TR-IMs. Entering all TRIM properties as simultaneous predictors of DMN-SC distance revealed a unique association with intrusiveness (*β* = −0.22, *t* = −2.16, *p* = 0.033), and no other properties, suggesting a degree of specificity between DMN-SC distance and the intrusiveness of TR-IMs.

**Figure 2.**
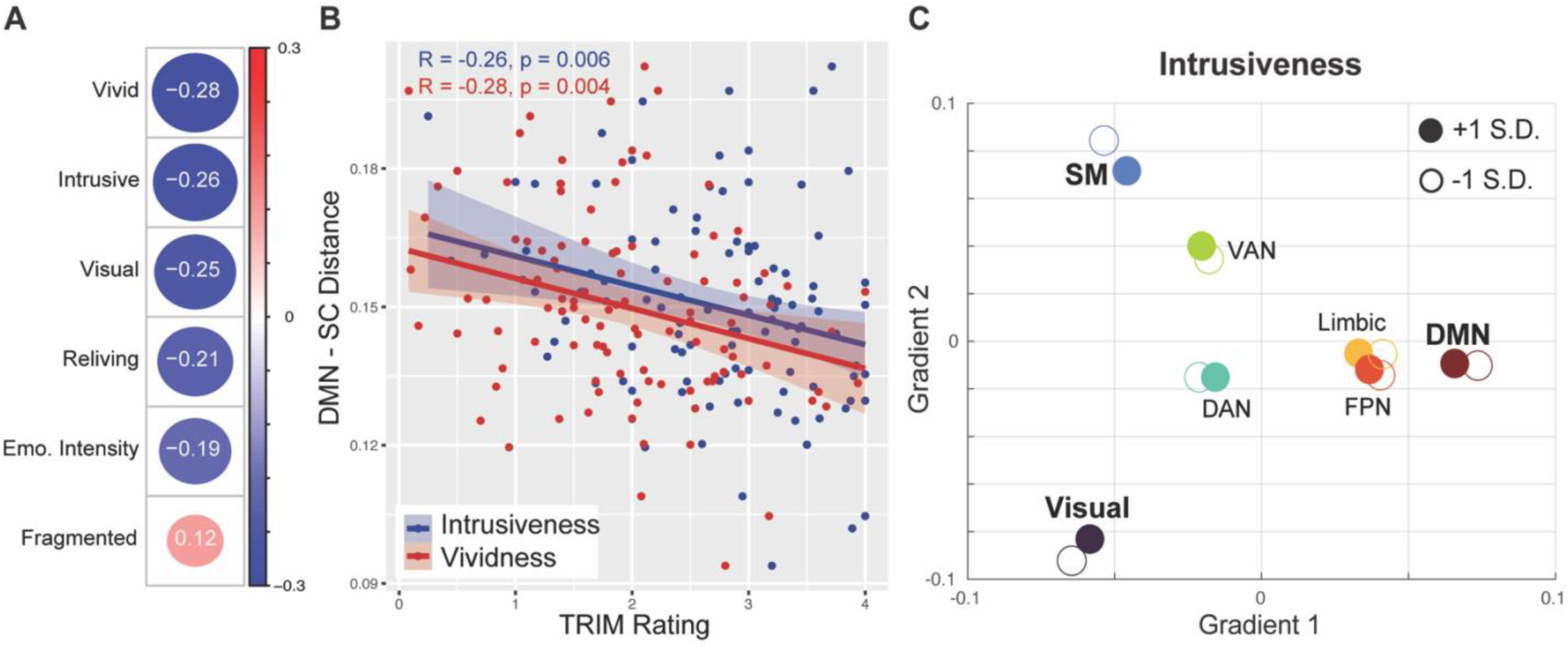
Associations between DMN-SC functional distance and TRIM properties. A) Pearson correlation coefficients demonstrating associations between greater DMN-SC distance and TRIM vividness and intrusiveness. There were additional associations seen with greater visual features and reliving properties of TRIMs; however, these did not survive correction for multiple comparisons. B) Scatterplots showing correlations between DMN-SC distance and TRIM intrusiveness (blue) and vividness (red). C) A schematic of Yeo’s 7 resting-state networks positioned within the 2D gradient space, color-coded by network, for individuals with high (+1 S.D.) and low (−1 S.D.) TRIM intrusiveness, demonstrating a compression of the Visual, SM, and DMN within the 2D gradient space for individuals with high TRIM intrusiveness. Solid circles represent participants with high TRIM intrusiveness while open circles represent those with low intrusiveness. SM = somatomotor network; DAN = dorsal attention network; VAN = ventral attention network; FPN = frontoparietal network; DMN = default mode network.

### DMN-SC distance was not associated with conventional clinical measures

Correlating Euclidean distance between the DMN and SC with conventional clinical assessments of posttraumatic stress symptoms revealed no association with total CAPS-5 PTSD symptom severity (*r* = 0.01, *p* = 0.937) or CAPS-5 Intrusive Reexperiencing symptoms (*r* = −0.06, *p* = 0.527). A Poisson regression model predicting the total number of TRIMs during the EMA period revealed no association between DMN-SC distance and TRIM frequency (*β* = −0.22, *z* = −0.25, p = 0.805).

Welch’s two sample t-test revealed no difference in DMN-SC distance between those with (M = 0.150, SD = 0.022) versus without PTSD (M = 0.149, SD = 0.014; *t(47.2)* = −0.23, *p* = 0.816). Moreover, PTSD diagnosis did not significantly moderate the associations between DMN-SC distance and TRIM vividness (*p* = 0.586) or intrusiveness (*p* = 0.125), suggesting these associations did not differ between participants with and without a PTSD diagnosis.

### Conventional resting-state functional connectivity

Correlating TRIM properties with conventional resting-state functional connectivity (rsFC) between the DMN and SC revealed an association between stronger DMN-SC connectivity and TRIM vividness (*r* = 0.27, *p* = 0.005) and a weak association with intrusiveness (*r* = 0.19, *p* = 0.044). Linear regression models entering DMN-SC distance and conventional rsFC as simultaneous predictors of TRIM properties revealed associations between vividness and both DMN-SC distance (*β* = −0.21, *t =* - 2.15*, p* = 0.034) and rsFC (*β* = 0.20, *t =* −2.00*, p* = 0.048). Conversely, for intrusiveness, an association was seen only with DMN-SC distance (*β* = −0.22, *t =* −2.24*, p* = 0.027), but not rsFC (*p* = 0.235). Notably, the estimated DMN-SC distance was modestly associated with conventional functional connectivity estimates between the DMN and SC (*r* = 0.34, *p* = 0.0003), indicating DMN-SC distance captures unique information beyond conventional rsFC. Overall, this suggests that gradient- based DMN-SC distance explains unique variance in TRIM vividness and intrusiveness, above and beyond conventional rsFC.

## DISCUSSION

In a symptomatic trauma-exposed sample of adults with frequent TRIMs, we found that the intrusiveness and vividness of TRIMs were associated with less functional segregation between the DMN and SC, as indexed by a shorter Euclidean distance in a two-dimensional space of functional connectivity gradients. When shared variance across TRIM properties was consideres, results suggested this association may be unique to intrusiveness and is independent of PTSD diagnosis or symptom severity. Moreover, our analyses suggest the DMN-SC distance metric explained unique variance in TRIM vividness and intrusiveness above and beyond standard rsFC measures. Together, these findings offer potential novel insights into the neural underpinnings of TRIMs, suggesting that the segregation of the DMN and SC within the hierarchical organization of the cortex may be particularly relevant to TRIM phenomenology.

Our metric of shorter functional distance between the DMN-SC can be interpreted as diminished functional segregation between these networks. Prior work utilizing the connectivity gradient method implemented here has characterized the hierarchical organization of the cortex, revealing maximal distance between the DMN and SC [35]. Functionally, this aligns with the respective roles of these opposing systems in cognition and perception. The SC supports rapid perception and processing of external sensory input, largely independent of higher-order cognitive processes that may bias or skew sensory perception. Conversely, the DMN supports high-level cognitive processes that arise from the complex integration of multimodal inputs and is thus largely removed from direct external sensory inputs that have not progressed through the cortical processing stream. Differences in intrinsic timescales of cortical regions across the cortical hierarchy further support this functional segregation: there is a progressive slowing of intrinsic activity along the cortical hierarchy, with the fastest intrinsic activity emerging within the SC and the slowest within the DMN [54–56]. This presumably allows for more rapid processing of external sensory information within the unimodal SC and slower, more refined processing within the DMN to enable the multimodal integration of multiple streams of information. Taken together, the maximal functional distance between the SC and DMN reflects the functional architecture of the cortex that allows the SC and DMN to have discrete functions at different levels of complexity and processing speed [54, 55, 57]. A shorter DMN-SC functional distance may therefore reflect weaker differentiation and an altered organizational architecture in which sensory perception is more closely coupled with and more readily enters higher-order cognitive processes, like ongoing thought and autobiographical memory.

The association we observed between shorter DMN-SC distance and greater TRIM vividness and intrusiveness aligns with this functional interpretation of a loss of DMN-SC segregation. Spontaneous autobiographical memory retrieval is associated with DMN activity [58] and the vividness of the retrieved memories tracks the degree to which the sensory cortex is reactivated [59, 60]. The engagement of higher-order cognitive networks like the DMN and minimal involvement of low-level sensory input in typical spontaneous memory retrieval may allow for efficient control and volitional suppression of the memories and thus minimize their attentional hijacking and intrusiveness [11, 61]. Conversely, TRIMs – marked by their vivid intrusiveness that makes them difficult to control – have been linked to less DMN activation, greater SC activation, and greater connectivity and coactivation between the DMN and SC [23, 31]. Recent work in trauma-exposed individuals and patients with PTSD further found that more frequent and stable brain states of coactivation between anticorrelated networks were linked to a shorter distance between such networks in gradient space [62]. This suggests that less functional segregation between the DMN and SC may be aligned with more frequent neural states of coactivation. Therefore, the observed loss of functional segregation between the DMN and SC may bias the brain towards states in which sensory representations of memories spontaneously intrude on conscious awareness through greater coactivation or shared information between the DMN and SC. This may explain the particularly robust association found with TRIM intrusiveness in the current study. Future studies combining connectivity gradient measures with measures of coactivation between networks may demonstrate the directional links between reduced DMN-SC segregation and greater DMN-SC coactivation in the context of the intruding trauma memories.

Notably, DMN-SC distance was not associated with conventional clinician-rated assessments of intrusive re-experiencing severity or the frequency of TRIMs during the EMA period. Rather than indexing the emergence of TRIMs or their clinical severity, DMN-SC segregation may instead offer insights into the neural correlates of their phenomenological experience. The emergence of TRIMs may involve classic PTSD-related circuitry, consisting of exaggerated amygdala activity, dysfunction of the hippocampus, and insufficient top-down control mechanisms governed by prefrontal cortices [63]. Nonetheless, the vivid and intrusive nature of TRIMs may serve as modifiable intermediate phenotypes that, if mitigated, may lead to changes in the phenomenology of TRIMs that make them more controllable and responsive to existing evidence-based treatments for PTSD [64].

We similarly found no associations with PTSD diagnosis – there were no differences in DMN- SC distance or its association with TRIM properties between those with versus without PTSD. This aligns with prior work showing a compression of the DMN and SC in gradient space in trauma exposed individuals, regardless of PTSD diagnosis, compared to non-trauma exposed controls [62]. Qualitative examination of network distances in gradient space revealed a compression of the DMN- SC distance in our symptomatic trauma exposed sample relative to the HCP reference dataset (Figure 1D). While quantitative comparisons are needed between trauma-exposed and non-trauma exposed adults to ascertain the influence of trauma on the observed DMN-SC segregation, our findings suggest that trauma exposure may be associated with a loss of DMN-SC segregation, particularly in individuals experiencing frequent TRIMs. Longitudinal sampling of trauma exposed individuals may further reveal the extent to which this loss of DMN-SC segregation is linked to the emergence of PTSD symptoms over time and the degree to which it is symptom specific.

Potential mechanisms underpinning this loss of functional segregation may lie in cortical inhibition. A balance of cortical excitation and inhibition serves as the foundation for the functional organization of the human cortex [65, 66]. This balance drives fluctuations in the integration and segregation of different cortical systems, resulting in intrinsic neural network dynamics. Recent work has demonstrated evidence for disinhibition of the sensory cortex in PTSD, indexed by deficits in alpha-frequency oscillations – a pattern of neural activity linked to the inhibition of cortical excitability [17, 67–69]. Additionally, alpha oscillations are implicated in the maintenance of resting-state neural networks through their role in long-range cortical communication [70–72] and increased resting-state alpha connectivity has been causally linked to stronger DMN connectivity [73]. Via posterior-to-frontal projections, alpha oscillations may regulate bottom-up information flow from the sensory cortex, gating the entry of sensory input into downstream processing [30, 74–76]. For example, a cascade of alpha suppression across the DMN has been observed during sensory cue-driven memory retrieval [77]. Notably, alpha deficits in patients with PTSD have been localized to both the visual cortex and DMN, including reduced alpha-frequency connectivity between and within these networks [30]. Taken together, alpha oscillations are positioned as potential candidates supporting the functional architecture of the cortex through cortical inhibition and may thus serve as a target for restoring optimal DMN-SC segregation. Future work utilizing neuromodulation techniques to increase alpha activity, such as transcranial alternating current stimulation [73, 78, 79] or alpha neurofeedback [67, 80, 81], may reveal a mechanistic link between alpha oscillations and DMN-SC segregation and generate novel neurotherapeutics for TRIMs.

The present study has several limitations to consider. While our focus on intrinsic, resting-state activity aligns with the spontaneous, “out of the blue” nature of TRIMs and provides important insights into the intrinsic functional organization of the brain that allows for such spontaneous intrusions, it precludes any claims about neural activity driving TRIMs. Future studies incorporating symptom provocation or capture paradigms may reveal patterns of neural activity that occur during the onset of TRIMs and how the intrinsic DMN-SC segregation contributes to these patterns of activity. Such studies could similarly address concerns about the time between the EMAs and neuroimaging in our study, as rsfMRI was performed days to weeks after completion of the EMA surveys. Measures of DMN-SC segregation during or more proximal to measures of TRIMs may reveal even more robust associations. Additionally, our sample was predominantly female which limits robust investigations into biological sex, which is particularly important given the demonstrated sex differences in PTSD. Future studies with a more balanced sex distribution may yield important insights into the role of sex and related biological systems in the neurobiological substrates of TRIMs.

In sum, greater vividness and intrusiveness of TRIMs in trauma-exposed adults were associated with a shorter functional distance between the SC and DMN. This suggests that less functional segregation of these intrinsically anticorrelated systems may contribute to the spontaneous and involuntary activation of sensory-based representations of trauma memories that intrude on ongoing cognition. These findings align with existing neurobiological models of TRIMs and PTSD, which implicate hyperactivity of sensory systems and dysfunction of autobiographical memory systems like the DMN. Moreover, these findings offer a parsimonious account for the phenomenological features of TRIMs, which involve both sensory-perceptual and cognitive-affective processes. Restoring the functional organization of these cortical systems could be a critical target for neurotherapeutics aimed at mitigating the distressing qualities of intrusive trauma memories.

## Data Availability

All data presented are available through the National Institute of Mental Health Data Archive, and are further available upon reasonable request to the senior author (IMR).

## Acknowledgments

This work was supported by NIH award R01-MH120400 (PI: IMR). KJC was supported by NIH award K23-MH137459 and a Brain and Behavior Foundation Young Investigator Award. IMR was partially supported by NIH award P50-MH115874 (Project 4 PIs – IMR, Scott L. Rauch; Program Directors – William A. Carlezon Jr. and Kerry J. Ressler). The authors would like to thank all participants for dedicating their time and energy to completing the study. We also thank the MRI technologists of the McLean Imaging Center.

